# Machine learning-based predictive models for perioperative major adverse cardiovascular events in patients with stable coronary artery disease undergoing non-cardiac surgery

**DOI:** 10.1101/2024.01.12.24301253

**Authors:** Liang Shen, YunPeng Jin, AXiang Pan, Kai Wang, RunZe Ye, YangKai Lin, Safraz Anwar, WeiCong Xia, Min Zhou, XiaoGang Guo

## Abstract

**Background:** Machine learning (ML)-based predictive models for perioperative major adverse cardiovascular events (MACEs) in patients with stable coronary artery disease (SCAD) undergoing non-cardiac surgery (NCS) have not been reported before.

**Methods:** Clinical data from 9171 consecutive adult patients with SCAD, who underwent NCS at the First Affiliated Hospital, Zhejiang University School of Medicine between January 2013 and May 2023, were used to develop and validate the prediction models. MACEs were defined as all-cause death, resuscitated cardiac arrest, myocardial infarction, heart failure and stroke perioperatively. Compare various resampling and feature selection methods to deal with data imbalance. A traditional logistic regression (the Revised Cardiac Risk index, RCRI) and nine ML models (logistic regression, support vector machine, Gaussian Naive Bayes, random forest, GBDT, XGBoost, LightGBM, CatBoost and best stacking ensemble model) were compared by the area under the receiver operating characteristic curve (AUROC) and the area under the precision recall curve (AUPRC). The calibration was assessed using the calibration curve and the patients’ net benefit was measured by decision curve analysis (DCA). Models were tested via 5-fold cross-validation. Feature importance was interpreted using SHapley Additive explanation (SHAP).

**Results:** Among 9171 patients, 514 (5.6%) developed MACEs. The XGBoost performed best in terms of AUROC (0.898) and AUPRC (0.479),which were better than the RCRI of AUROC (0.716) and AUPRC (0.185), Delong test and Permutation test P<0.001, respectively. The calibration curve of XGBoost performance accurately predicted the risk of MACEs (brier score 0.040), the DCA results showed that the XGBoost had a high net benefit for predicting MACEs. The top-ranked stacking ensemble model consisting of CatBoost, GBDT, GNB, and LR proved to be the best, with an AUROC value of 0.894 (95% CI 0.860-0.928) and an AUPRC value of 0.485 (95% CI 0.383-0.587). Using the mean absolute SHAP values, we identified the top 20 important features.

**Conclusion:** The first ML-based perioperative MACEs prediction models for patients with SCAD were successfully developed and validated. High-risk patients for MACEs can be effectively identified and targeted interventions can be made to reduce the incidence of MACEs.

**Lay Summary:** We performed a retrospective machine learning classification study of MACEs in patients with SCAD undergoing non-cardiac surgery to develop and validate an optimal prediction model. In this study, we analyzed the data missing mechanism and identified the best missing data interpolation method, while applying appropriate resampling techniques and feature selection methods for data imbalance characteristics, and ultimately identified 24 preoperative features for building a machine learning predictive model. Eight independent machine learning prediction models and stacking ensemble models were built, and the models were evaluated comprehensively using ROC curve, PRC curve, calibration plots and DCA curve.

- We have adopted a series of widely used machine learning algorithms and model evaluation techniques to build clinical prediction models, and achieved better performance and clinical practicability than the classical RCRI model, which has taken the first step to explore the research in this field.
- The prediction results based on the optimal machine learning model are interpretable, output the importance ranking and impact degree of the top 20 features of MACEs risk prediction, and are consistent with clinical interpretation, which is conducive to the application of the model in clinical practice.

## Background

Millions of patients undergo non-cardiac surgeries (NCS) worldwide every year [1], more than 18% of them accompany with stable coronary artery disease (SCAD) [2]. Perioperative major adverse cardiovascular events (MACEs) occur in 5.7–10.0% patients with SCAD undergoing NCS [3], which exceeded significantly compared to only 2.5–3.0% MACEs occurrence rate in the general population [4]. MACEs represented a significant source of perioperative morbidity and mortality, including cardiac arrest, myocardial infarction (MI), heart failure (HF) and stroke [5]. Accordingly, it is important to evaluate the risk of MACEs for patients with SCAD undergoing NCS.

Current guidelines highly recommended the use of predictive models to assess the risk of perioperative MACEs [6, 7]. The most commonly used models are the Revised Cardiac Risk index (RCRI) [8]. The RCRI is simple and widely validated worldwide. However, recent large cohort studies have suggested that the RCRI does not have a strong discriminatory ability [9], especially in patients with known SCAD [10].

Machine learning (ML) is an area of artificial intelligence (AI) where algorithms are employed for identification of patterns in datasets, and have demonstrated superior predictive performance on nonlinear data as compared to conventional linear models such as logistic or cox regression [11]. It learns from the data, as opposed to regression which stems from theory and assumptions, benefiting from human intervention and subject knowledge to specify a model [12].As a result, the application of innovative machine-learning techniques capable of capturing nonlinearity in clinical practice is imperative.

The objective of this study was to derive and validate a ML model based on easily acquired preoperative clinical data, that can predict perioperative MACEs in patients with SCAD scheduled for NCS. As far as we know, such a prediction model has not previously been reported.

## Methods

### Study design and population

We performed a retrospective machine learning classification study (outcomes were binary categorical) of MACEs in patients with SCAD undergoing NCS to develop (train) and validate (test) an optimal prediction model. The study design route flowchart is shown in Figure 1. The machine learning model predicts a future diagnosis of perioperative MACEs based on features obtained from preoperative usual clinical care, including demographics, previous diseases, surgical information, preoperative electrocardiogram (ECG), preoperative echocardiography and preoperative laboratory tests results such as hemoglobin levels. This study used data from 9,171 adult patients with SCAD who underwent NCS at the First Affiliated Hospital, Zhejiang University School of Medicine (FAHZU) between January 2013 and May 2023.

**Figure 1.**
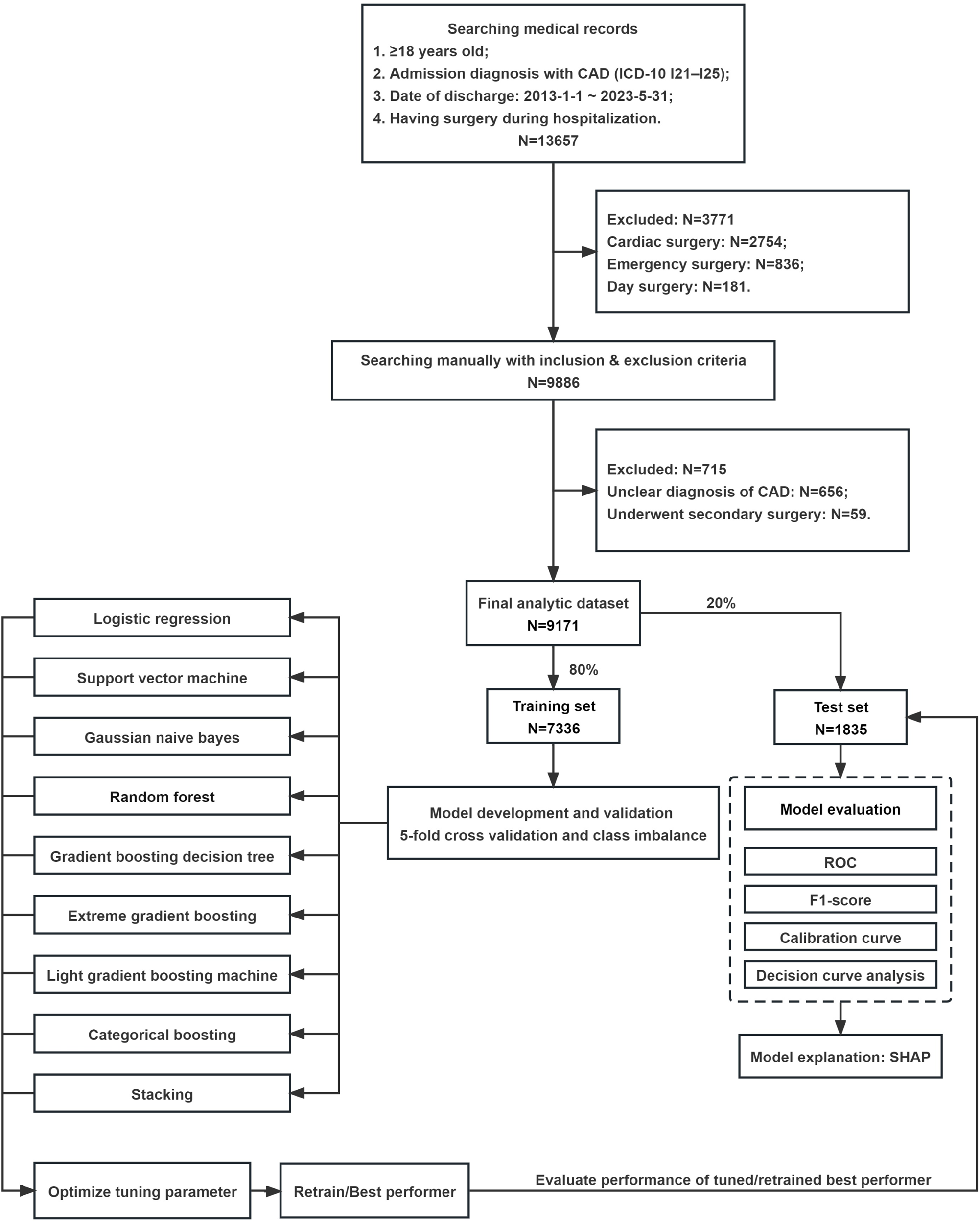
Flowchart of study design route.

This study was conducted according to Transparent Reporting of Multivariable Prediction Models for Individual Prognosis or Diagnosis (TRIPOD) and “Guidelines for Development and Reporting Machine-Learning Predictive Models in Biomedical Research: A Multidisciplinary View”. It complied with the principles of the Declaration of Helsinki and was approved by the Institutional Ethics Review Committee of the FAHZU (No. of ethical approval: IIT20230114A). Written informed consent was waived owing to the nature of the retrospective study design and the collected data was managed in a de-identified form. This study was executed and reported in accordance with STrengthening the Reporting of OBservational studies in Epidemiology (STROBE) guidelines.

### Inclusion and exclusion criteria

We extracted the study dataset of patients aged 18 years and older who were hospitalized for surgery with previous SCAD between January 1, 2013 and May 31, 2023 from FAHZU’s clinical data warehouse. The types of surgery were elective NCS based on the American College of Cardiology (ACC)/American Heart Association (AHA) guidelines of perioperative cardiovascular evaluation [13]. SCAD was diagnosed if any of the following conditions were met: angiographic demonstration of coronary stenosis >50%, history of MI (>3 months before enrolment), history of coronary revascularization (>3 months before enrolment), positive myocardial perfusion scintigraphy, positive exercise stress test, or typical symptoms of angina pectoris with simultaneous signs of myocardial ischemia on the ECG [14]. We excluded people who underwent cardiac surgery, emergency surgery, day surgery, and people who underwent multiple surgeries during a single hospital stay. All patients were evaluated by routine preoperative assessment.

### Data collection and preprocessing

The electronic medical record system in FAHZU was used in this study. The International Classification of Diseases, Tenth Edition (ICD-10) has been used to extract the target population. We identified all discharges over 10 years from the surgery department with a diagnosis of CAD. Further manual screening of medical records was performed according to the inclusion/exclusion criteria. We then listed all available clinical data from the electronic medical record system and performed feature selection. The study omitted variables with a high rate of missing values (e.g., hs-CRP and troponin I). Finally, a total of 64 pre-operative variables were collected, including patients’ demographics (e.g. age, sex and Body Mass Index (BMI)), pre-existing diseases (e.g. MI, HF, hypertension and diabetes), surgical information (e.g. surgical type, duration of surgery (DOS) and general anesthesia (GA)), preoperative ECG (e.g. abnormal Q waves (AQW) and ST-T wave abnormalities (ST-Ta)), preoperative echocardiography (e.g. left ventricular ejection fraction(LVEF), regional wall motion abnormality (RWMA), left ventricle diastolic dysfunction (LVDD) and pulmonary hypertension (PH)), pre-operative laboratory parameters (e.g. Hemoglobin (Hb), Fasting blood glucose (FBG) and Creatinine (Scr)), pre-operative drugs (e.g. Nitrates and Insulin), American Society of Anesthesiologists Physical Status (ASA PS). The putative predictors were chosen on the basis of previous studies and the clinical experiences of the investigators.

For each feature, we calculated the missing rate in the training dataset, analyzed the missingness mechanisms, and then selected the appropriate missing value data imputation method according to the missingness mechanisms. Additionally, standardization is essential for ensuring that all feature values are on the same scale and assigned the same weight. All continuous variables (e.g., BMI, laboratory values) were scaled using StandardScaler or MinMaxScaler in the Scikit-learn package, the classification of non-binary variables (e.g., surgical type) were one-hot encoded, and the variables with ordinal characteristics (e.g., ASA PS) were coded with the ordinal encoder.

### Outcomes (Study endpoints and definitions)

The primary outcome was a composite of MACEs (all-cause death, resuscitated cardiac arrest, MI, HF and stroke) intraoperatively or during hospitalization postoperatively. Cardiac arrest was defined as the loss of circulation prompting resuscitation requiring chest compressions, defibrillation, or both [15]. MI was defined as acute myocardial injury with clinical evidence of acute myocardial ischemia [16]. Troponin levels were not routinely checked on all enrolled patients. They were ordered based on routine clinical practice whenever the treating physician suspected MI based on the clinical status of the patient or ECG findings. HF was diagnosed mainly by active clinical symptoms or physical examination findings of dyspnea, orthopnea, peripheral edema, jugular venous distention, rales, third heart sound, or chest x-ray with pulmonary vascular redistribution or pulmonary edema [17]. Stroke was diagnosed by a neurology consultant based on new neurological findings that were confirmed by imaging studies [18].

### Class imbalance

The data set of this study included 8657 negative samples (majority class) and 514 positive cases (minority class), with an imbalance ratio (IR) of 16.84:1, indicating a serious class imbalance. Most standard machine learning algorithms assume or expect that classification problems have balanced class distributions of equal costs. As a result, these algorithms are not efficient at handling the complex and imbalanced data sets that are prevalent in the real world, especially in the medical field. Solving class imbalance data is mainly realized from two levels of data and algorithm [19]. In this study, resampling and feature selection are mainly used to process research data, and ensemble learning models are compared to explore the most appropriate methods to deal with data imbalance. In order to comprehensively analyze the classification performance of imbalanced data sets, area under the receiver operating characteristic curve (AUROC) and area under the precision and recall curve (AUPRC) are emphasized in model evaluation.

### Resampling for class imbalance

Resampling is a technique to balance a dataset by reducing the number of majority classes or increasing the number of minority classes. Among them, Synthetic minority over-sampling technique (SMOTE) [20] represents the most widely used method among the resampling methods. Overfitting caused by random oversampling can be effectively overcome by interpolating new synthetic instances in the line between some minority samples and their k-nearest neighbors. The adaptive synthetic (ADASYN) [21] sampling method is to use a weighted distribution for different minority class examples according to their level of difficulty in learning, and minority samples that are more difficult to learn will generate more synthetic data. The SMOTE+EEN [22] hybrid sampling method uses Edited Nearest Neighbors (ENN) technology to clean up overlapping samples after the SMOTE algorithm generates a new synthetic dataset. In this study, SMOTE, ADASYN and SMOTE+ENN sampling methods were used to resample the training set data. Then, we controlled the sampling strategy so that the ratio of positive samples to negative samples in the resampling dataset is 1. Finally, we trained the eXtreme Gradient Boosting (XGBoost) model with resampling data combined with cross-validation, and compared the model’s performance metrics on the internal validation set. We used correlation functions in the Python library imbalance-learn to implement resampling.

### Feature selection for class imbalance

Feature selection is also a feasible technique to deal with imbalanced classification problems. More representative feature sets are selected to remove irrelevant and redundant features, thereby improving classification performance and efficiency. Feature selection is carried out on imbalanced data to optimize the feature space, find a space that tends to represent concepts of a few classes, and then correct the classifier’s bias towards the majority classes. This study contrasts four widely used feature selection methods based on different strategies, including the correlation-based feature selection (CFS) algorithm [23], Boruta algorithm [24], BorutaShap algorithm [25], and recursive feature elimination (RFE) [26]. CFS algorithm is a multivariate filter method that chooses subsets of features that themselves are uncorrelated but show high correlation with the class, independent of any learning method, and successfully applied to mortality prediction in three-vessel disease [27]. Boruta algorithm is to compare the importance of the real predictor variables with those of random so-called shadow variables using statistical testing and several runs of XGBoost algorithm. BorutaShap algorithm is an extension of the Boruta algorithm that leverages the SHapley Additive explanation (SHAP) value as a measure of feature importance with XGBoost classifier. The RFE algorithm starts with a base model built on all features. A specific proportion of the least important features are then removed and a new base model is generated using the remaining features. These steps are recursively applied until a single feature is left as input. In this study, XGBoost, Light Gradient Boosting Machine (LightGBM), Random Forest (RF), support vector machine (SVM) and logistic regression (LR) were selected as the base models for RFE. Feature selection takes XGBoost model as performance evaluation. Based on data resampling, AUROC performance is evaluated on validation set through cross-validation combined with automatic hyperparameter optimization, and the optimal performance is taken as input feature set of machine learning model.

### Machine learning model development

We used random stratification to divide the data set into a training dataset (80%) and a test dataset (20%). Stratification ensured that the proportions of the cases in the training datasets and test datasets were equal, which improved the stability of the model. The training dataset was used for model building, and the test dataset was used as a hold-out dataset for external validation and did not participate in model development (including data balancing processing) and hyperparameter selection. We used randomly stratified 5-fold cross-validation combined with optimal resampling strategy on the training data set to adjust the hyperparameters in the model and output the internal validation performance, which can avoid overfitting and assess the stability of the models. After obtaining the optimal hyperparameters, we used the model developed in the training dataset for performance evaluation on the hold-out testing dataset. In order to achieve the best prediction, eight independent models were built for this study, including LR, SVM, Gaussian Naïve Bayesian (GNB), RF, gradient boosting decision tree (GBDT), XGBoost, LightGBM, and categorical boosting (CatBoost). Tree based ensemble models have been applied to other clinical tasks with excellent performance compared to traditional machine learning algorithms [28]. Each model provides the same input variables that are optimally selected based on feature selection, and in order to avoid collinearity between variables affecting the performance of the prediction model, multicollinearity and correlation analysis are performed on the optimally selected samples before modeling. We adjusted the hyperparameters during the model building process based on the Optuna optimization library of Bayesian optimization [29], where the optimized measure is the AUROC. Finally, based on 8 independently optimized machine learning prediction models, we further used the stacking ensemble model, which has been proven to be superior to independent machine learning in many fields [30, 31].

### Machine learning model evaluation

We developed the models using the training dataset performed 1000 rounds of bootstrapping on the hold-out testing dataset to report results. We reported numerical results for accuracy, precision, recall, F1 score, AUROC, and AUPRC. To evaluate the overall performance, we plotted receiver-operating characteristic (ROC) curves and precision–recall curves (PRC). The ROC is the ratio of sensitivity to (1-specificity). According to the AUROC evaluation of model performance, models with a larger AUROC are considered to have better performance. On the other hand, the PRC illustrates the trade-off between recall (sensitivity) and precision (positive predictive value). Models with high performance tend to have a balance of high recall and precision, yielding large AUPRC values. The statistical comparison of AUROC values and AURPC values were each computed using Delong Test and Permutation test [32, 33]. A calibration plot was used to evaluate the agreement between the observed and expected values based on the probability of perioperative MACEs predicted by various models, and calculated the calibration metrics of the Brier score [34]. The clinical application value of decision curve analysis (DCA) evaluation model.

### Comparison with RCRI

To determine whether the new developed models in our study would improve upon discrimination of cardiovascular risk prediction, we also developed a baseline model that mimics the classical clinical scoring system RCRI [8]. The baseline model was a logistic regression model that included only RCRI. Our newly developed machine learning model was compared numerically and statistically with this baseline model in AUROC and AUPRC performance.

### Machine learning model explainability

We analyze and visualize the feature importance of the generated predictive model to comprehend how the model makes predictions and realize an explainable machine learning model. We used SHAP to analyze and visualize the effect of feature importance on perioperative MACEs risk based on best-performing predictive models. The SHAP value represents the effect of features on the prediction in terms of direction and range by calculating a weighted average and marginal distribution, which is calculated by comparing the predicted differences in all possible combinations containing and withholding each feature.

### Statistical analysis and modeling tools

The normality of the distribution of continuous variables was tested using the Shapiro–Wilk test. Normally distributed continuous variables were expressed as mean ± standard deviation (SD) and compared using the independent samples t-test. Skewed continuous variables were expressed as median and interquartile range (IQR) and compared using the Mann–Whitney U-test. Categorical variables are expressed as frequencies and percentages and using chi-square tests or Fisher’s exact probability tests. The differences were considered to be statistically significant at p < 0.05. Machine learning model development and evaluation was performed in python 3.6 using scikit-learn packages.

## Results

### Participant characteristics

We eliminated 4,486 patients based on exclusion criteria, and ultimately 9,171 patients were included in our study. Among them, 514 (5.6%) patients suffered perioperative MACEs, as shown in Supplementary Table 1. Table 1 presents baseline clinical characteristics of the training and testing sets, respectively, and univariate analyses with and without MACEs. Overall, the baseline clinical characteristics of the training set and testing set samples appeared to be similar. Patients underwent a wide range of surgeries as expected in a tertiary referral hospital with a median age of 70 (IQR, 64–77) years. General abdominal, thoracic, and vascular surgeries were most often performed.

**Table 1.**
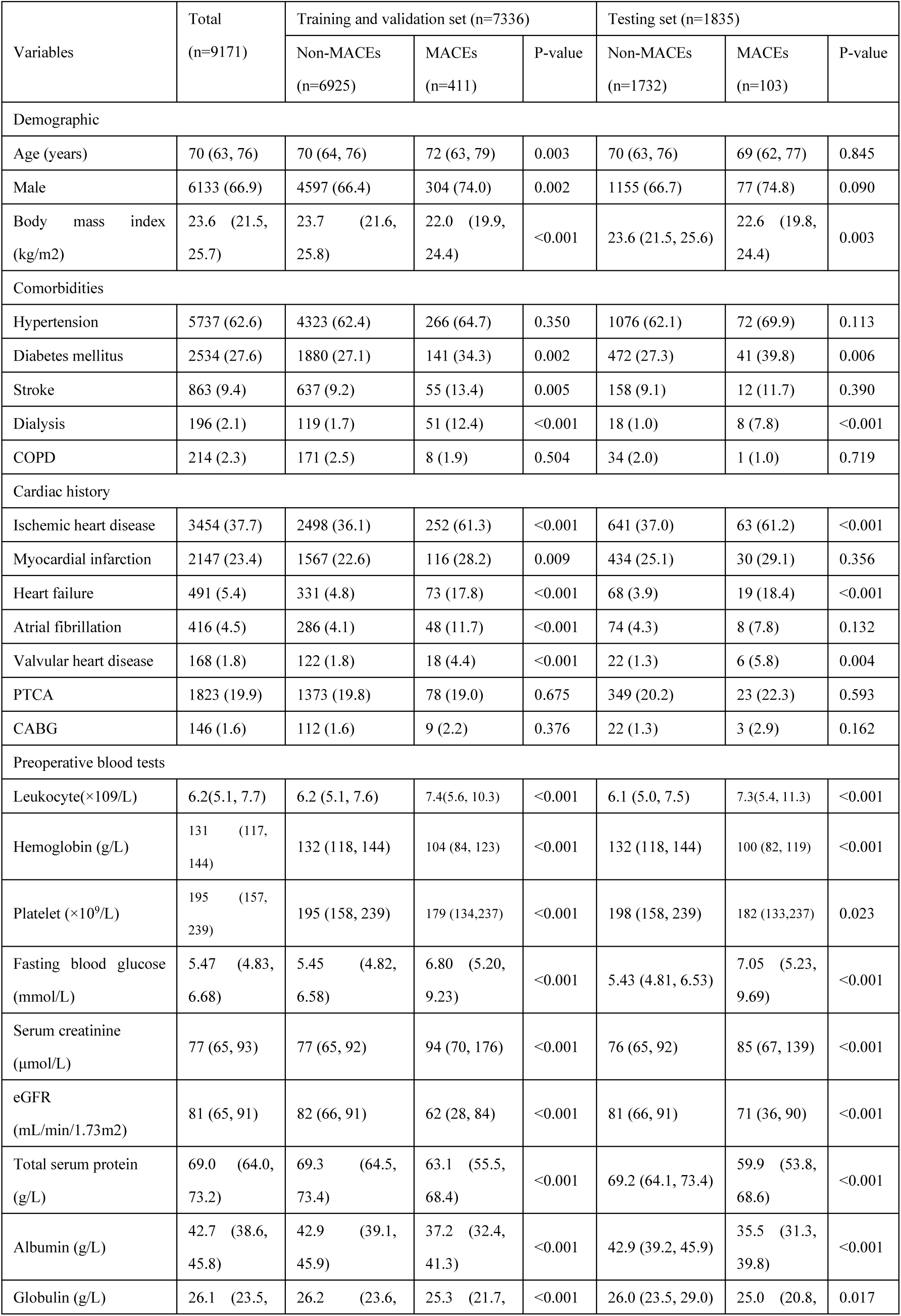
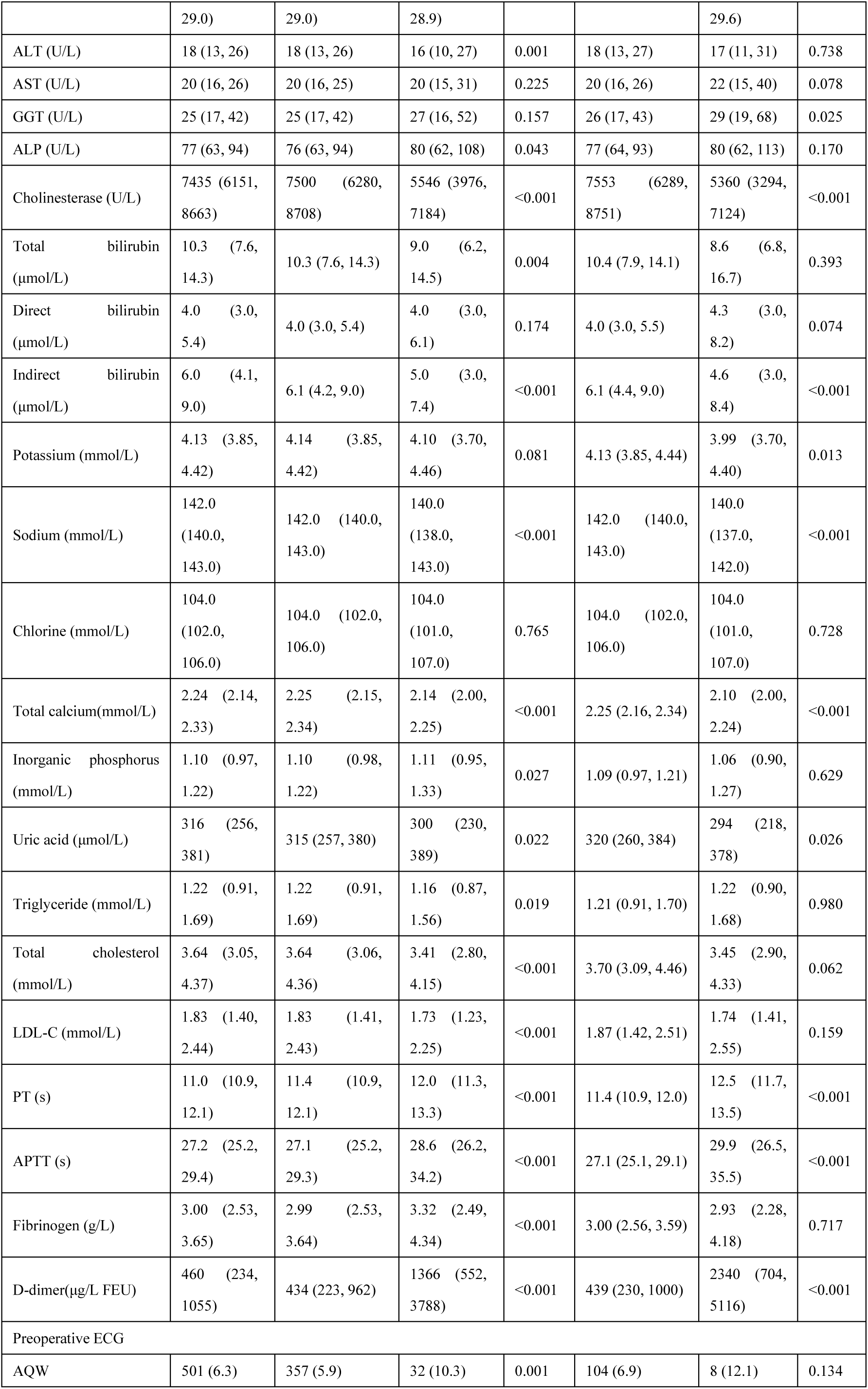
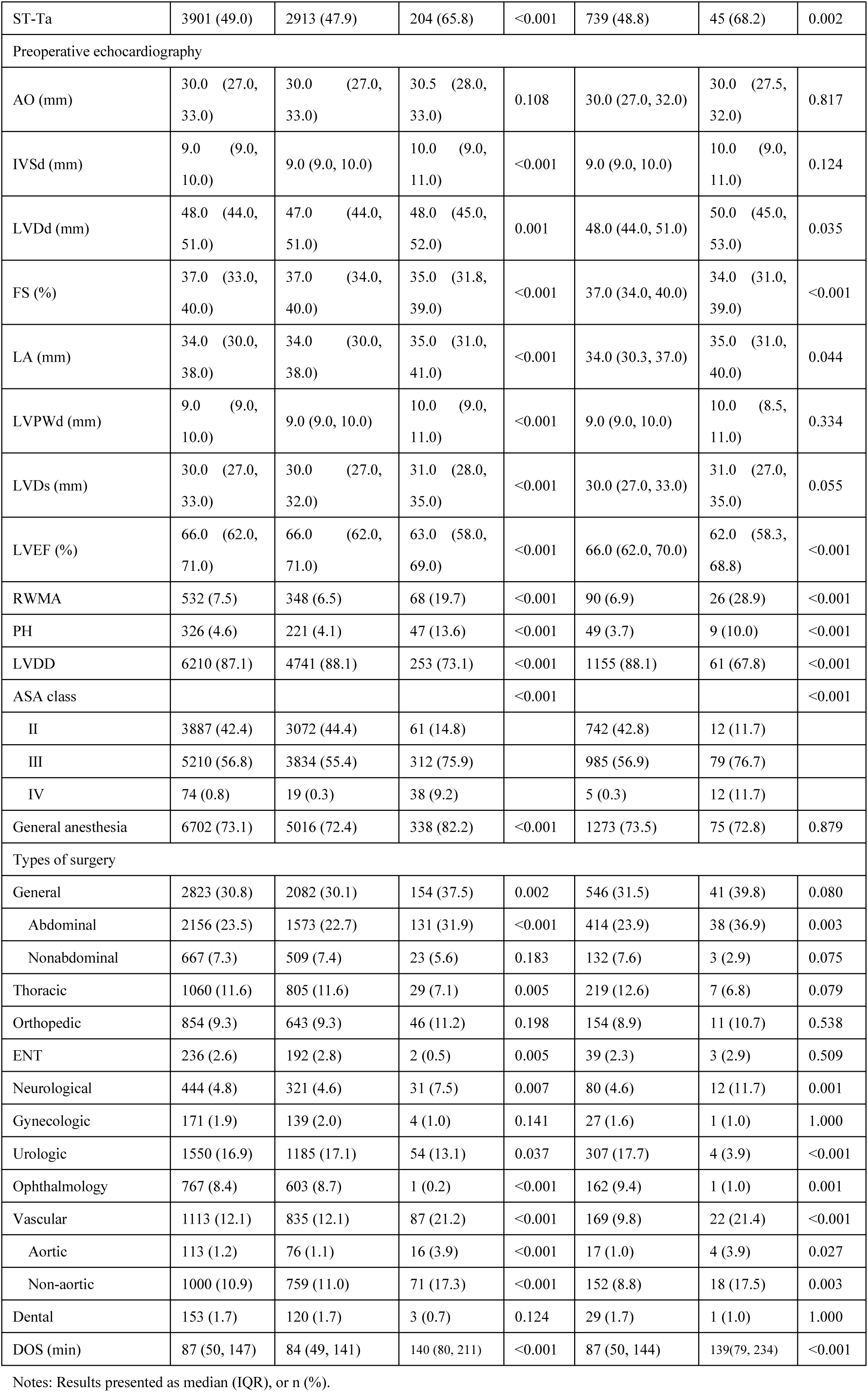
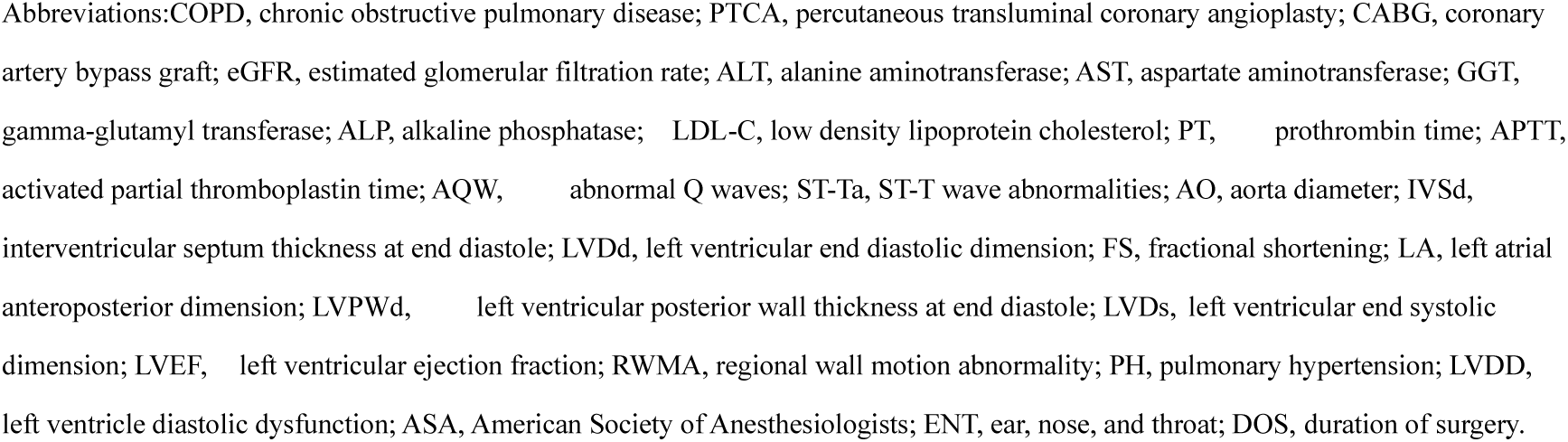
Baseline clinical characteristics of the study population and their association with perioperative outcomes.

### Missing-value characteristics

The average proportion of missing values in this study dataset was 7.69%, and the proportion of the training and validation datasets (7.62%) and the testing dataset (7.99%) were basically the same (Supplementary Table 2). The missing values of preoperative laboratory tests, preoperative ECG and preoperative echocardiography were mainly due to the fact that the patient did not complete the corresponding examination in FAHZU, while the missing BMI value is mainly due to the patient’s body being unable to measure normally. At the same time, we tested the correlation of missing values of different variables (Supplementary Figure. 1a), the correlation between the variables measured as a companion test (e.g., laboratory, ECG and echocardiogram tests) was high (absolute correlation value ≥ 0.7), while the correlation between BMI, laboratory variables, ECG variables, echocardiogram variables, and DOS variables was not remarkable (absolute correlation value ≤ 0.2). Comparison of data missing distribution and completeness of variables (Supplementary Figure. 1b, 1c), missing values include categorical and continuous variables, and there is no uniform pattern of missing values for each variable. Based on the miss at random mechanism of variables, we compared a variety of missing value imputation algorithm on the XGBoost model, cross-validation on the training dataset (Supplementary Table 3), and finally selected the k-Nearest Neighbor (KNN) imputation algorithm with the best performance in this study.

### Resample method

The imbalanced training set data is processed by resampling method to overcome the performance loss caused by data imbalance. Table 2 presents the internal verification results of LR, RF and XGBoost models in each training set. As shown in the table, although the three models also achieved high AUROC and AUPRC performance before balancing the data, the extremely high specificity and extremely low sensitivity indicated that the classification model without data balance could not well identify MACEs patients (minority class) due to the inter-class imbalance in the data. In contrast, after using SMOTE, ANSYN and SMOTE+ENN for data balance, the sensitivity of the three models has significantly improved, and the indicators of AUROC and AUPRC have also increased. The results showed that the data balancing processing can effectively improve the recognition performance of the classification model for the few class samples. Further contrast the same prediction model based on different resampling methods to observe the corresponding changes in specificity and sensitivity. SMOTE+ENN is the best for data balancing and will be applied to model development and evaluation.

**Table 2.** The internal verification results of models trained were obtained by using different balanced class methods combined with cross-validation using all features.

| Models | Methods | AUROC(95%CI) | AUPRC(95%CI) | Accuracy | Specificity | Sensitivity | Youden Index |
| --- | --- | --- | --- | --- | --- | --- | --- |
| LR | - | 0.879(+/-0.024) | 0.451(+/-0.126) | 0.879 | 0.991 | 0.270 | 0.261 |
| RF | - | 0.864(+/-0.036) | 0.414(+/-0.095) | 0.944 | 1.000 | 0.000 | 0.000 |
| XGBoost | - | 0.862(+/-0.047) | 0.403(+/-0.055) | 0.946 | 1.000 | 0.034 | 0.034 |
| LR | SMOTE | 0.867(+/-0.036) | 0.412(+/-0.121) | 0.824 | 0.828 | 0.757 | 0.585 |
| RF | SMOTE | 0.870(+/-0.047) | 0.386(+/-0.105) | 0.943 | 0.979 | 0.343 | 0.322 |
| XGBoost | SMOTE | 0.875(+/-0.041) | 0.427(+/-0.137) | 0.942 | 0.976 | 0.372 | 0.348 |
| LR | ANSYN | 0.864(+/-0.041) | 0.408(+/-0.134) | 0.806 | 0.809 | 0.764 | 0.573 |
| RF | ANSYN | 0.871(+/-0.044) | 0.388(+/-0.104) | 0.941 | 0.973 | 0.399 | 0.372 |
| XGBoost | ANSYN | 0.877(+/-0.035) | 0.415(+/-0.137) | 0.947 | 0.982 | 0.348 | 0.330 |
| LR | SMOTE+ENN | 0.868(+/-0.036) | 0.396(+/-0.105) | 0.726 | 0.719 | 0.842 | 0.561 |
| RF | SMOTE+ENN | 0.863(+/-0.053) | 0.360(+/-0.096) | 0.864 | 0.691 | 0.874 | 0.565 |
| XGBoost | SMOTE+ENN | 0.872(+/-0.037) | 0.397(+/-0.105) | 0.894 | 0.907 | 0.672 | 0.579 |

### Feature selection

The total number of features in the data set after the pre-processing is 75, and the features are analyzed and selected sequentially based on eight feature selection methods. Figure 2A corresponds to the optimal feature subset of the eight feature selection methods and the AUROC performance of the internal validation set after cross-validation on the training set. The performance of the optimal subset after feature selection is better than that of the full feature (Supplementary Table 4), but it is also related to the selected feature subset, in which the performance of the feature subset selected by RFE-XGB and RFE-LR methods is the best. Figure 2B shows that RFE combines five kinds of basis learning models, selects the best subset recursively based on the feature importance ranking of the learning model, and evaluating the AUROC performance of the internal validation set after cross-validation on the training set. When RFE-XGB and RFE-RF are used for feature selection, AUROC performance is maintained at a relatively high level when the number of features of the optimal subset is greater than or equal to 3, and the average AUROC is greater than 8.0.

**Figure 2.**
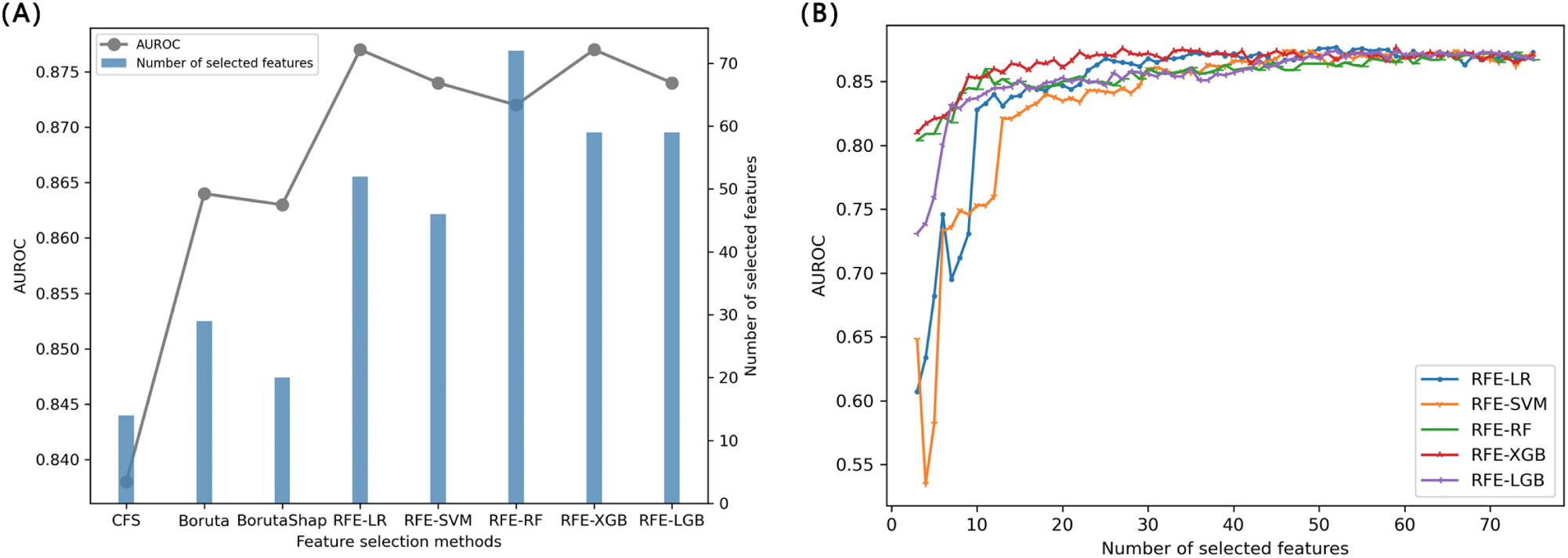
Performance assessment of feature selection methods. (A) The optimal feature subset of the eight feature selection methods and the AUROC performance of the internal validation set after cross-validation on the training set. (B) RFE combines five kinds of basis learning models, selects the best subset recursively based on the feature importance ranking of the learning model, and evaluating the AUROC performance of the internal validation set after cross-validation on the training set.

The performance and interpretability of the model are fully balanced, and the number threshold of features contained in the optimal feature subset is controlled between 3 and 30, and the feature combination with the best performance is selected by comparing eight feature selection methods. These features included patients’ demographics [BMI], pre-existing diseases [Ischemic heart disease (IHD), and Dialysis], surgical information [DOS], preoperative echocardiography [fractional shortening (FS), left ventricular end systolic dimension (LVDs), LVEF and RWMA), pre-operative laboratory parameters [Leukocyte, Hb, FBF, Scr, Estimated glomerular filtration rate (eGFR), Total serum protein (TSP), Albumin (ALB), AST, Cholinesterase (ChE), Total bilirubin (TB), Total calcium (tCa), Chlorine, APTT, Fibrinogen (FB) and D-dimer], ASA PS. Subsequently, we carried out multicollinearity analysis of the selected features. First, correlation coefficients and corresponding P-values of the features were drawn in the heat map, and it was found that TSP and ALB, Scr, eGFR and Dialysis may have collinearity (Supplementary Figure 2). The variance inflation factor (VIF) was further used for the feature multicollinearity test, and VIF values less than 5 indicated weak multicollinearity (Supplementary Table 5). It indicates that the features selected in this study can effectively avoid the negative effects of feature collinearity on the classification performance of the model. We ultimately used the above 24 features for model development and evaluation.

### Machine learning model performance

Eight independent candidate models were constructed for perioperative MACEs prediction using the twenty-four variables mentioned above. Figure 3 A, B presents the AUROC and the AUPRC of each candidate modeling method in test-set data. All eight candidate models exhibited superior prediction performances in terms of AUROC and AUPRC, compared to that of the Baseline-RCRI model, with significant differences in both AUROC and AUPRC by using the Delong Test and Permutation test (P <0.001). The XGBoost method delivered the best performance in terms of AUROC (0.898) and AUPRC (0.479). Table 3 presents the other metrics of mean values of bootstrapping performance of each model. Further, the DCA curves (Figure 3C) demonstrate that the eight candidate models exhibited a greater net benefit along with the threshold probability compared with Baseline-RCRI models. The calibration curve of the eight candidate models is closer to the curve with a slope of 45° than the Baseline-RCRI model, indicating the better accuracy (Figure 3D), while the Brier scores were calculated, none exceeding 0.04, and was better than RCRI (0.05). Machine learning model hyperparameters are listed in Supplementary Table 6.

**Figure 3.**
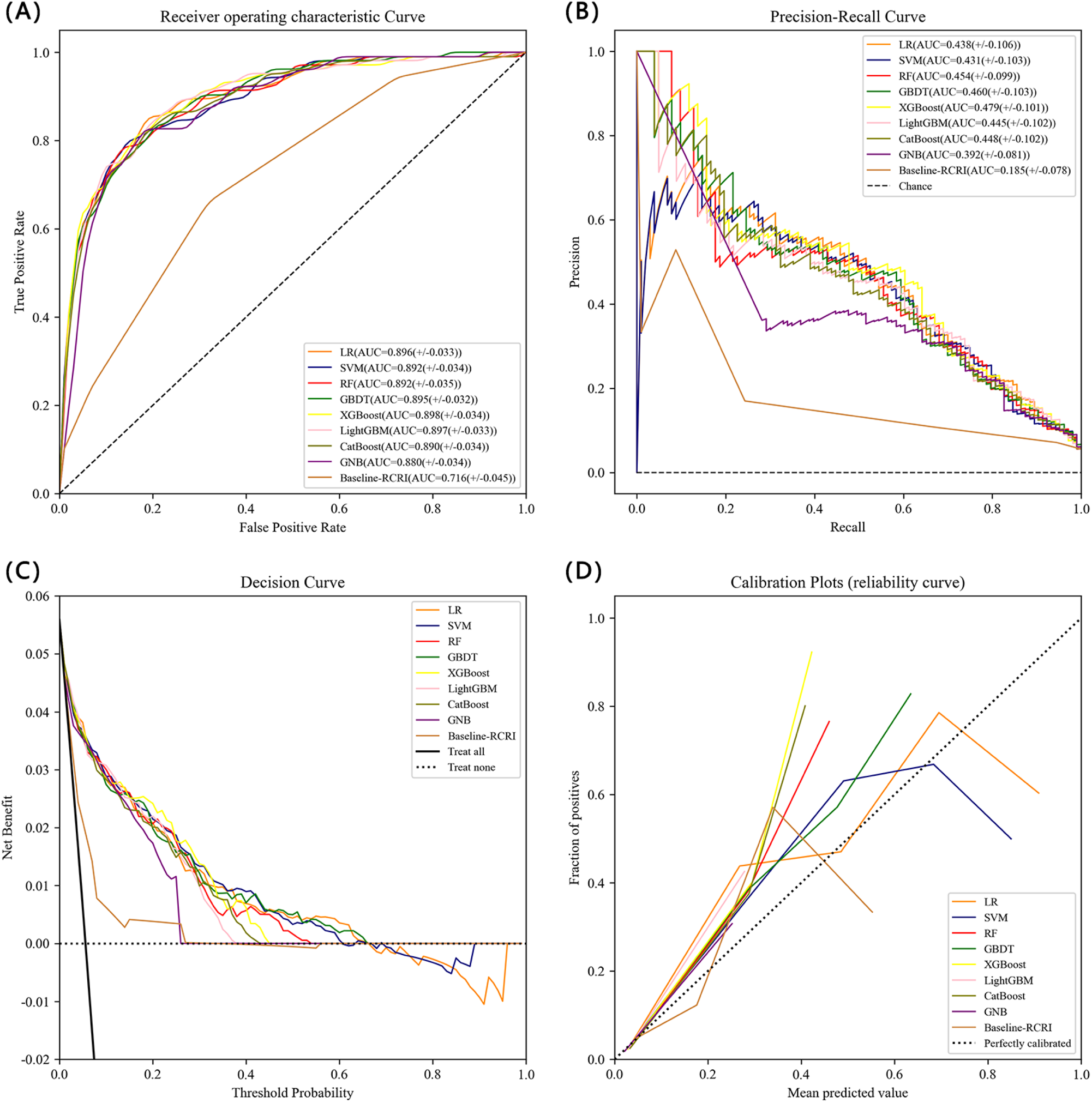
Performance assessment of the models. (A) Receiver operating characteristic curve (ROC) of MACEs prediction models in testing set. (B) Precision-Recall curve (PRC) of MACEs prediction models in testing set. (C) Decision curve analysis (DCA) for the nine MACEs prediction models in the testing set. (D) Calibration plots of MACEs prediction models in the testing set.

**Table 3.**
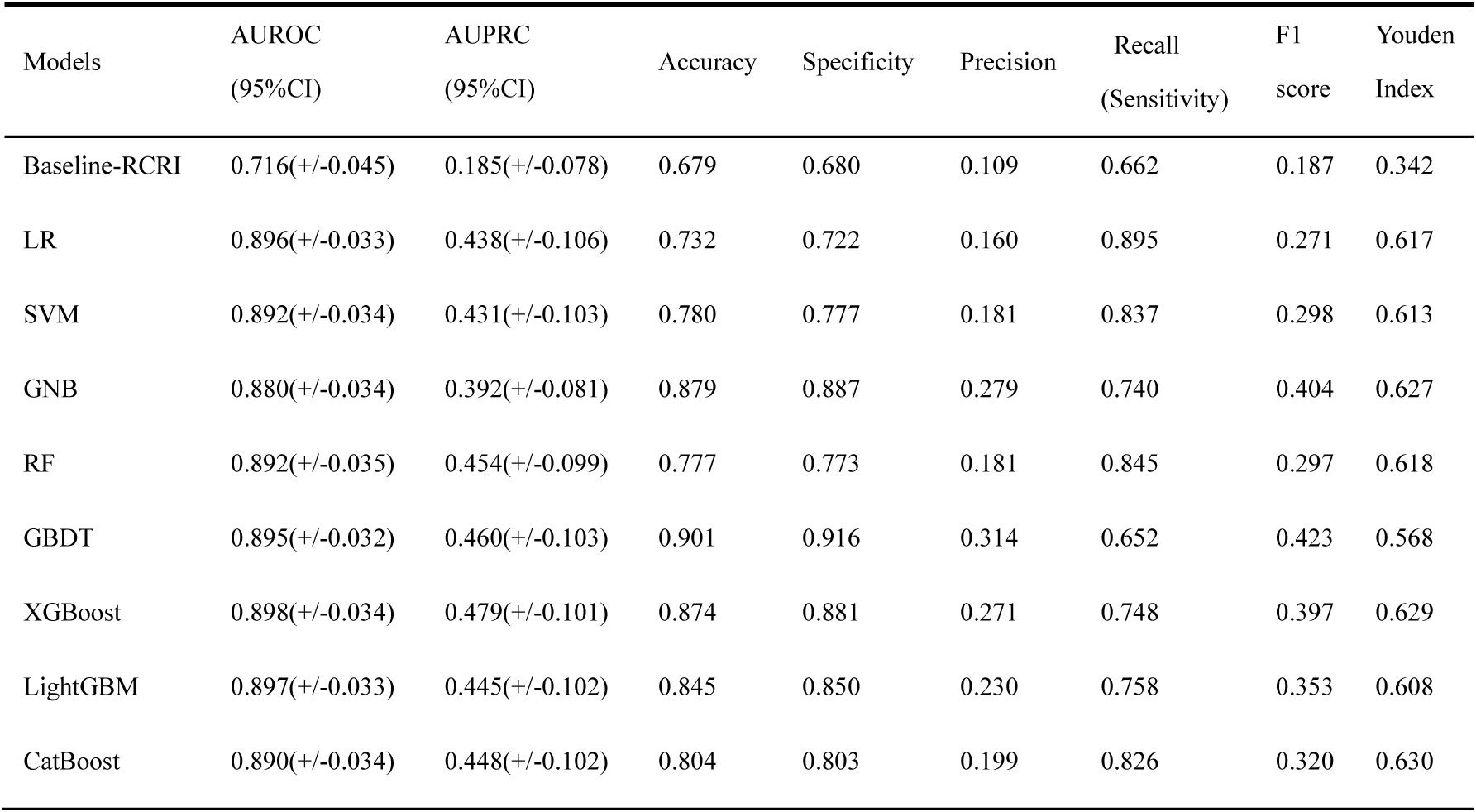
The predictive performance in the test set of the 9 models. LR: logistic regression; SVM: support vector machine; GNB: Gaussian Naive Bayes; RF: random forest; GBDT: gradient boosting decision tree; XGBoost: extreme gradient boosting; LightGBM: light gradient boosting machine; CatBoost: categorical boosting; AUROC: area under the receiver operating characteristic curve; AUPRC: area under the precision recall curve.

The eight prediction models were developed based on the training set data and 5-fold cross-validation. The AUROC and AUPRC per fold on the internal validation set of the prediction models are shown in Supplementary Table 7. The results show that the performance is similar in each fold of the verification set, which indicates that the prediction model has good stability. Further, the internal verification results of different prediction models were compared with the external verification results. Figure 4 shows the comparison results of AUROC (A) and AUPRC (B) performance indicators of the verification set and the test set. The results show that the comprehensive performance of the validation set and the test set of the prediction model is similar, and the performance of the test set is slightly better than that of the verification set, which indicates that various prediction models in this study have good generalization performance.

**Figure 4.**
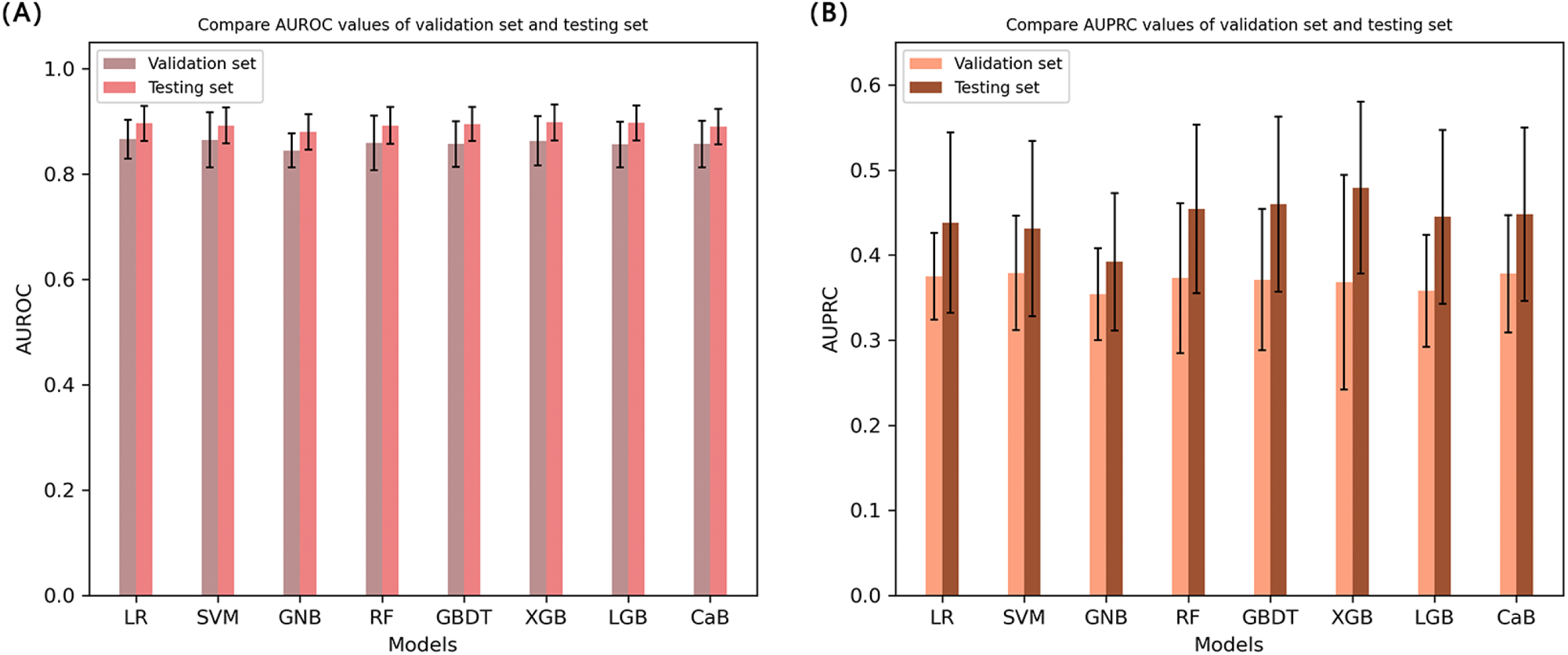
Compare the performance of validation sets and testing sets on different models. (A) Compare AUROC values of validation set and testing set. (B) Compare AUPRC values of validation set and testing set.

### Model explainability

Based on the optimal independent model XGBoost, by using SHAP analysis (Figure 5 A, B), we determined the top 20 features including IHD, ASA PS, Hb, DOS, LVDs, D-dimer, ALB, Chlorine, FBG, ChE, Leukocyte, Scr, RWMA, eGFR, BMI, TSP, APTT, tCa, Dialysis, and LVEF as important features for predicting new onset MACEs. In Figure 5A, we presented the relationships between their values and the effect of the model output. Intuitively, IHD, ASA PS, DOS, LVDs, D-dimer, FBG, Leukocyte, Scr, RWMA, APTT, and Dialysis were positively correlated with the MACEs, whereas Hb, ALB, Chlorine, ChE, eGFR, BMI, TSP, tCa, and LVEF were negatively correlated with the MACEs.

**Figure 5.**
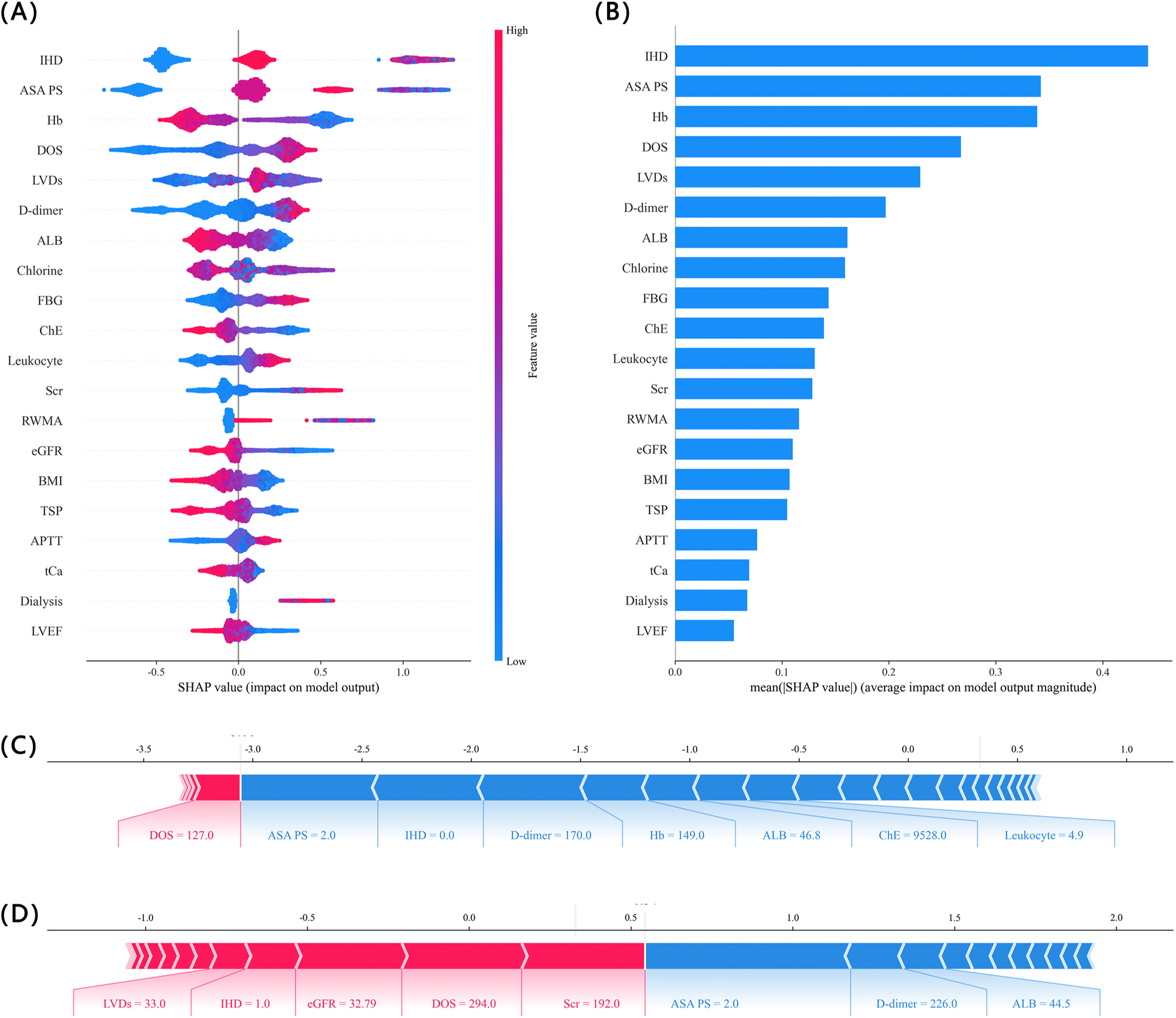
SHAP interprets the XGBoost Predictive model. (A) The SHAP analysis was performed on the XGBoost. Each row of the graph represents a variable and the horizontal coordinate is the SHAP value, which represents the distribution of the effect of the variable on the risk of MACEs, with positive values indicating a risk of MACEs and negative values indicating no risk of MACEs. A point represents a patient, while red represents a higher value and blue represents a lower value. (B) The average of the absolute values of the SHAP values for each variable in the XGBoost is taken as the significance of that variable. (C) Examples of negative predictions for MACEs. (D) Examples of positive predictions for MACEs.

In addition to the overall effect, we applied the SHAP framework to explain individual cases by providing influential features. Figure 5 shows 2 examples of random selection - a negative prediction (C) and a positive prediction (D). Features in blue represent features that contribute to a lower risk while features in red will push up the risk. These visualizations give users detailed information about how the model makes predictions and allow them to make appropriate interventions before the new onset MACEs.

### Model Stacking

Stacking ensemble models were subsequently developed, and the stacking ensemble model is a two-layer structure, the first layer is composed of multiple base models, and the second layer is fixed as a logistic regression model. Based on the 8 independent models of this study, 247 model combinations were listed by exhaustive method, and then the stacking model and output performance indicators were established and sorted. Overall, the top-ranked stacked ensemble model consisting of CatBoost, GBDT, GNB, and LR proved to be the best, with an AUROC value of 0.894 (95% CI 0.860-0.928) and an AUPRC value of 0.485 (95% CI 0.383-0.587). Compared to the independent optimal prediction model XGBoost, the stacking model showed slightly higher AUPRC performance and net benefit value (Figure 6), as well as higher sensitivity (0.788).

**Figure 6.**
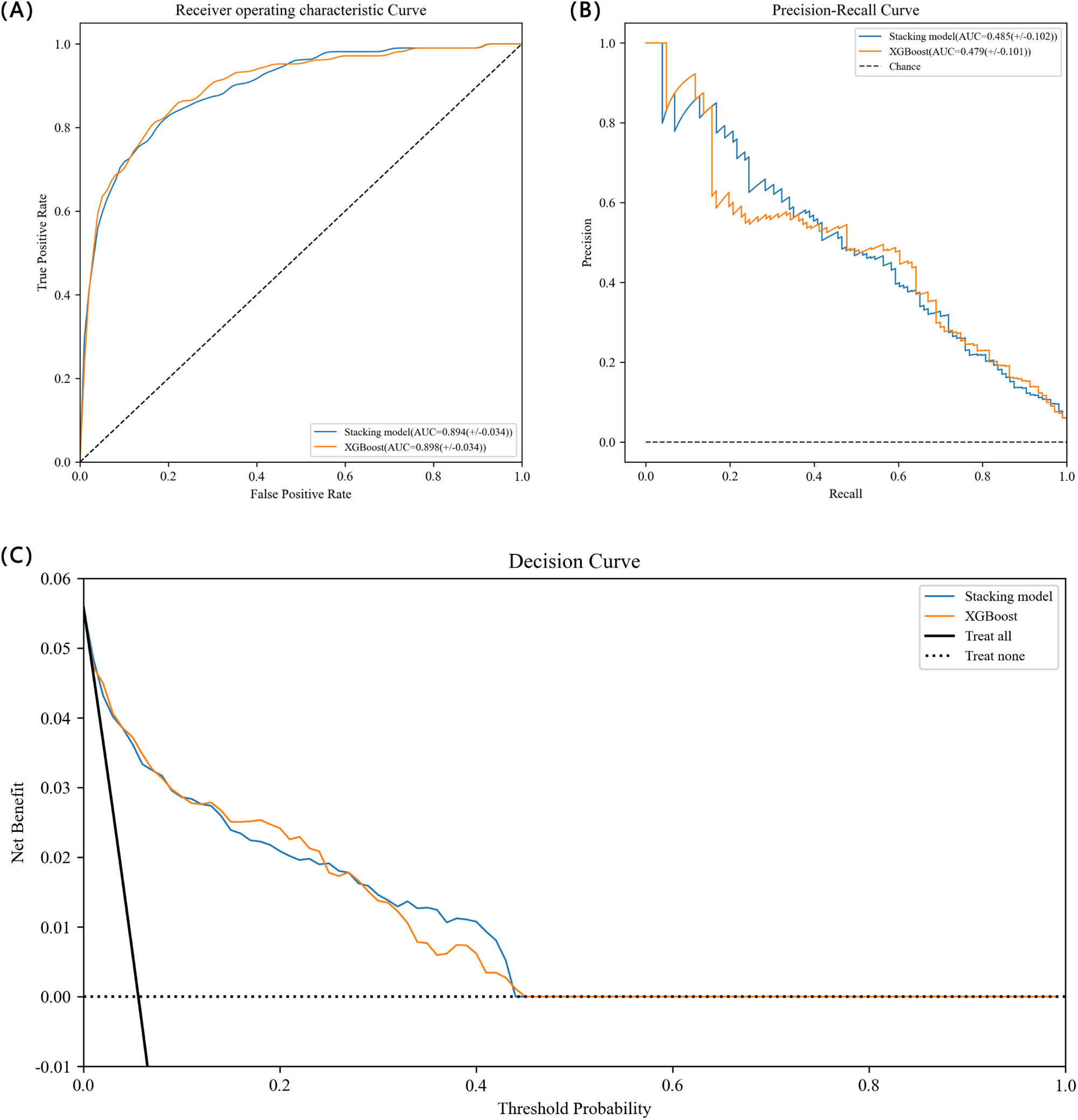
The ROC, PR and DCA curve performance of stacking model. ROC: receiver operating characteristic; PR: precision-recall; DCA: decision curve analysis; AUROC: area under the receiver operating characteristic curve; AUPRC: area under precision-recall curve.

## Discussion

To our knowledge, this is the first study of a systematic framework based on machine learning techniques to develop multiple models, evaluate performance, and select the highest-performing model to predict perioperative MACEs in patients with SCAD scheduled for NCS. This study shows that compared with the classical prediction model RCRI [8], the eight independent machine learning prediction models and the optimal stacking ensemble model have a great improvement in performance and clinical utility, and have satisfactory generalization, among which XGBoost has the best performance in the independent machine learning prediction model. IHD, ASA PS, Hb, DOS, LVDs, D-dimer, ALB, Chlorine, FBG and ChE were the top 10 important characteristics affecting model performance. In addition, the model combining CatBoost, GBDT, GNB, and LR is considered to be the best model for stacking ensemble learning, further improving the AUPRC, clinical utility, and sensitivity of the predictive model. These findings help to identify perioperative MACEs risk in patients with SCAD scheduled for NCS before surgery and to provide targeted clinical care through timely intervention.

From the data set used in this study, there are some inherent characteristics in the data set that affect the classification performance of the prediction model, such as data missing and data imbalance. This paper proposes a series of research strategies to improve the performance of machine learning classification algorithms based on data processing. Missing data is a common occurrence in clinical research, and improper processing will significantly affect the efficacy of the classification model [35]. First, the identification of missing data mechanism is the basis of selecting missing data imputation method. There are three typical mechanisms causing missing data: missing completely at random (MCAR), missing at random (MAR), and missing not at random (MNAR). In this study, the missing data mechanism was determined to be MAR through correlation test and integrity comparison of missing data. On this basis, different missing data imputation methods were compared, and the most effective one was selected according to the performance output of the internal validation set, while directly deleting missing data would lead to estimation bias [36]. Second, in terms of data balance, this study reconstructs the data set from the data itself. Primarily, resampling method is used to optimize the sample space, and then feature selection method is combined to optimize the feature space. After resampling the unbalanced data, the performance of all models has been significantly improved, especially the sensitivity of RF model increased from 0 to 0.874 after the SMOTE+ENN method. SMOTE+ENN method, after SMOTE algorithm generates new synthetic data set, uses ENN clearing technology to reduce the problem that SMOTE often introduces more noise and overfitting to some extent [37]. On the basis of selecting suitable resampling methods, the feature selection method based on AUC evaluation criteria was adopted in this study. Some studies have concluded through experiments that feature selection is more important than classification method selection in order to overcome overfitting problems and achieve better classification performance [38]. The feature selection method is used to delete category-irrelevant features, reduce the dimension of data, find a space that tends to represent the concepts of minority classes, correct the classifier’s bias to the majority classes, and solve the unbalanced classification problem with poor classification performance of minority classes [39]. Although univariate analysis of clinical features was performed in this study, threshold filtering features were not directly set according to p-value in the process of feature selection, mainly considering that univariate analysis might ignore the interaction between features [40]. The 8 feature selection methods in this study mainly select features based on the relevance and importance of features, and the results show that the performance of the model after feature selection is indeed improved. The internal verification performance comparison of different feature selection methods finally determines 24 effective features. The distribution of feature categories was fairly balanced, including 1 feature of patients’ demographics, 2 features of pre-existing diseases, 1 feature of surgical information, 4 features of preoperative echocardiography, 15 features of pre-operative laboratory parameters and ASA PS. The feature subset after feature selection is continued to be multicollinear detected and processed by correlation coefficient test and VIF method, so as to reduce the complexity of classification model construction and improve model stability and generalization ability.

Considering the clinical applicability of the prediction model, the prediction model in this study uses only routine clinical and laboratory data and selects only a small number of features, which is conducive to the data being automatically collected through the program and the model being applied to other institutions to obtain stable performance. In addition, this study only used preoperative data, not intraoperative data, so it has the ability to predict prognosis before surgery. The use of intraoperative data may improve prediction accuracy but may lead to an exaggeration of model performance and delay in implementing interventions to patients.

In our study, XGBoost provided the best predictive performance among the independent models built. Compared with the classical model RCRI, which uses logistic regression with 6 equal weight variables, the main advantages of XGBoost model are the ability to capture the nonlinear relationships between the model features and the outcome, as well as having higher order interactions between features. Evaluates the performance of the machine learning predict model in external test set, performs 1000 rounds of bootstrapping sampling method to report the result confidence interval to evaluate the stability of the model parameters. The ROC curve and its corresponding AUC are a function of the sensitivity and specificity of the predictive model and are used to quantify the overall ability of the test to correctly identify normal and abnormal ones. The prediction models developed in this study all had AUROC values greater than 0.88 on the test set, and the AUROC value of the XGBoost model was close to 0.9, which means that on average, the test will correctly predict abnormal outcomes 90% of the time, and the model has excellent prediction ability. Compared with the ROC curve, the PRC curve assesses the true proportion of the positive prediction and provides more information on the prediction assessment of the imbalanced dataset. The AUPRC value is low compared to the higher AUROC value, and the AUPRC value of the prediction model we developed is between 0.39 and 0.48, but it is still much higher than the classical model RCRI (AUPRC=0.185). The reason for the low AUPRC value is that the low incidence of perioperative MACEs leads to the imbalance of research data categories, although we have performed techniques such as resampling and feature selection before model construction to try to reduce the impact of data imbalance on model performance, it is difficult to significantly increase the AUPRC value. In addition to the evaluation of the predictive performance of the model, we also reported the calibration of the model, which was reported in the form of a calibration plot. The calibration plot shows that the predictive model developed in this study is well calibrated, although it appears to have a tendency to slightly underestimate the risk of MACEs. ROC curve, PRC curve, sensitivity, specificity, and calibration for assessing predictive models are reported, but do not provide answers as to whether models are effective in clinical practice. Decision analysis attempts to address the question of clinical utility assessment by combining the clinical outcomes of the model [41]. The DCA curve shows that the developed prediction model has good clinical practicability and has obvious advantages over the classical RCRI model. Finally, in order to further improve the performance of the prediction model, we used a stacking ensemble algorithm based on the optimization of independent machine learning prediction models [31]. Compared with the XGBoost model, the optimal stacking model combining CatBoost, GBDT, GNB and LR improved the AUPRC, clinical practicability and sensitivity of the prediction model.

The interpretability of machine learning predictions requires attention, so that doctors can understand them, trust them and gain useful insights for the clinical practice. However, the “black box” nature of the ML algorithm and the difficulty for clinicians to understand and trust the interpretation of the data are still the most difficult hurdle to overcome [42]. XGBoost was excellent at predicting post-operative mortality, with performance comparable to deep learning [11]. Compared to deep learning, XGBoost has the advantage of using the SHAP to interpret the model output, demonstrating the possibility of solving the “black box” problem. In this study, taking XGBoost as an example, we calculated the SHAP values of important features and used SHAP graphs to intuitively show the impact of features on the prediction model. IHD, ASA PS, and Hb were the top three important features of the XGBoost. This is consistent with clinical practice because IHD, ASA PS, and Hb have been used as important predictive indicators in previous clinical prediction models [43]. It is worth mentioning that, chlorine was not considered as a predictive indicator in univariate analysis. However, chlorine was the eighth important feature in XGBoost. In fact, chlorine has been proved to be related to heart failure [44]. Most of the predictive indicators in XGBoost can be improved, which means the patients can benefit from appropriate preoperative intervention. This study had several strengths. To begin with, there is a higher incidence of perioperative MACEs in the patient with SCAD scheduled for NCS compared to the general population, but few studies have been conducted. We have adopted a series of widely used machine learning algorithms and model evaluation techniques to build clinical prediction models, and achieved better performance and clinical practicability than the classical RCRI model, which has taken the first step to explore the research in this field. In addition, the prediction results based on the optimal machine learning model are interpretable, output the importance ranking and impact degree of the top 20 features of MACEs risk prediction, and are consistent with clinical interpretation, which is conducive to the application of the model in clinical practice. Moreover, we use Bayesian algorithm to automatically adjust the model hyperparameters, so that the selection of appropriate missing data imputation method, resampling technology and feature selection method can be combined with automatic hyperparameter tuning, and it is also confirmed that the appropriate resampling technology combined with feature selection can greatly improve the impact of data imbalance on model performance. Finally, we put forward the stacking ensemble model, and use the exhaustion method to form 247 stacking models, evaluate the performance of each model in turn, and select the optimal stacking model. Compared with the optimal independent model XGBoost, the optimal stacking ensemble model showed slightly higher AUPRC performance and clinical utility, with higher sensitivity.

The study has several limitations. First, in terms of feature collection, currently features are mainly from single text data of electronic medical records, and high-quality features can be extracted based on image (such as electrocardiogram) recognition technology. Second, we developed models based on data sets from a single medical center. Exploring the predictability of this model in other medical centers could add even more value. However, it should be noted that the data set in this study was extracted from 10 years of data from a large medical center with multiple hospitals, and the model evaluation used a testing set that was completely independent of model development as external validation. Third, as a retrospective study, the effect of predicting perioperative MACEs risk on prognosis in patients with SCAD remains unknown.

In future studies, we will further develop and validate current machine learning models based on data from other large, multicentre populations that can predict different types of MACEs (e.g., all-cause death, resuscitated cardiac arrest, MI, HF and stroke) and provide interventions accordingly. Another direction is to integrate the model into the clinician’s workflow by designing an interactive interface, integrating with electronic medical record systems, and further exploring the model’s impact on clinician behavior and patient outcomes.

## Conclusion

In this study, we analyzed the data missing mechanism and identified the best missing data interpolation method, while applying appropriate resampling techniques and feature selection methods for data imbalance characteristics, and ultimately identified 24 preoperative features for building a machine learning predictive model. Eight independent machine learning prediction models and stacking ensemble models were built, and the models were evaluated comprehensively using ROC curve, PRC curve, calibration plots and DCA curve. The results show that the machine learning prediction model developed in this study has better prediction performance and generalization than the classical RCRI model, and has the potential to be applied in clinical practice. With further validation and refinement, machine learning predictive models can help more effectively assess perioperative MACEs risk and target interventions to at-risk populations, as well as provide better clinical access and ease of use.

## List of abbreviations

NCS: non-cardiac surgery
SCAD: stable coronary artery disease
MACEs: major adverse cardiovascular events
MI: myocardial infarction
HF: heart failure
RCRI: Revised Cardiac Risk index
ML: Machine learning
AI: artificial intelligence
ECG: electrocardiogram
FAHZU: First Affiliated Hospital, Zhejiang University School of Medicine
TRIPOD: Transparent Reporting of Multivariable Prediction Models for Individual Prognosis or Diagnosis
STROBE: STrengthening the Reporting of OBservational studies in Epidemiology
ACC: American College of Cardiology
AHA: American Heart Association
ICD-10: International Classification of Diseases, Tenth Edition
BMI: Body Mass Index
DOS: duration of surgery
GA: general anesthesia
AQW: abnormal Q waves
ST-Ta: ST-T wave abnormalities
LVEF: left ventricular ejection fraction
RWMA: regional wall motion abnormality
LVDD: left ventricle diastolic dysfunction
PH: pulmonary hypertension
Hb: Hemoglobin
FBG: Fasting blood glucose
Scr: Creatinine
ASA PS: American Society of Anesthesiologists Physical Status
IR: imbalance ratio
AUROC: area under the receiver operating characteristic curve
AUPRC: area under the precision and recall curve
SMOTE: Synthetic minority over-sampling technique
ADASYN: adaptive synthetic
ENN: Edited Nearest Neighbors
XGBoost: eXtreme Gradient Boosting
CFS: correlation-based feature selection
RFE: recursive feature elimination
SHAP: SHapley Additive exPlanation
LightGBM: Light Gradient Boosting Machine
RF: Random Forest
LR: logistic regression
SVM: support vector machine
GNB: Gaussian Naïve Bayesian
GBDT: gradient boosting decision tree
CatBoost: categorical boosting
ROC: receiver-operating characteristic
PRC: curves and precision–recall curves
DCA: decision curve analysis
SD: standard deviation
IQR: interquartile range
KNN: k-Nearest Neighbor
IHD: Ischemic heart disease
FS: fractional shortening
LVDs: left ventricular end systolic dimension
eGFR: Estimated glomerular filtration rate
TSP: Total serum protein
ALB: Albumin
ChE: Cholinesterase
TB: Total bilirubin
tCa: Total calcium
FB: Fibrinogen
VIF: variance inflation factor
MCAR: missing completely at random
MAR: missing at random
MNAR: missing not at random

## Declarations

### Availability of data and materials

The datasets generated during and/or analyzed during the current study are not publicly available but are available from the corresponding author at reasonable request.

### Ethics approval and consent to participate

This study was approved by the Institutional Ethics Review Committee of the the First Affiliated Hospital, Zhejiang University School of Medicine (No. of ethical approval: IIT20230114A). Written informed consent was waived owing to the nature of the retrospective study design and the collected data was managed in a de-identified form.

### Consent for publication

Not applicable.

### Authors’ contributions

LS and YPJ contributed to the conception, design, analysed data and coding of the work, and drafted the manuscript. AXP, KW, RZY, YKL, SA and WCX contributed to the acquisition, analysis, and interpretation of data for the work. MZ and XGG were responsible for conceptualization and formal analysis, critically revised the manuscript. All authors read and approved the final manuscript.

## Acknowledgments

This work was supported by grants from the National Natural Science Foundation of China, Joint Funds from the National Natural Science Foundation of China, and grants from the Key Research and Development Plan of Zhejiang Province

## Supplementary Material

data supplement.docx

